# cardiovascular safety of COVID-19 vaccines in real-world studies: a systematic review and meta-analysis

**DOI:** 10.1101/2022.08.01.22278252

**Authors:** Yafei Chang, Guoli Lv, Chao Liu, Bin Luo, Erwen Huang

## Abstract

**Aims:** To assess the association between COVID-19 vaccines and the risk of major adverse cardiovascular events (MACE) in the real world and to provide a reliable evidence-based basis for the cardiovascular safety of COVID-19 vaccines.

**Methods:** We conducted a comprehensive search in databases from January 1, 2020 to June 15, 2022 for observational studies, that included reporting of MACE and COVID-19 vaccines were included. Random-effects or fixed-effects models were used to estimate the pooled incidence rate and risk ratio of MACE after vaccination. Meta-regression, subgroup analyses, publication bias, sensitivity analyses were performed to evaluate the process and quality of meta-analysis.

**Results:** The analyses included data from 43 studies reporting of 16,978 cases, 28,451 cases, and 96,269 cases of myocarditis, myocardial infarction, and cardiac arrhythmia, respectively. The overall incidence rate was 14.8 events per million persons of myocarditis, and 1.73 and 9.6 events per 10,000 persons of myocardial infarction and cardiac arrhythmia after COVID-19 vaccination, respectively. Overall and subgroup analyses showed the increased risks of myocarditis associated with second dose (RR, 2.09; 95%CI: 1.59-2.58), third dose (RR, 2.02; 95%CI: 1.40-2.91), mRNA-1273 (RR, 3.13; 95%CI: 2.11-4.14), or BNT162b2 (RR, 1.57; 95%CI: 1.30-1.85) vaccination. The risk ratios of myocarditis events were more frequently in males than in females (3.44, 2.61-4.54), in younger than in older (2.20, 1.06-4.55). No significant increase risk of myocardial infarction (RR, 0.96; 95%CI: 0.84-1.08) or cardiac arrhythmia (RR, 0.98; 95%CI: 0.84-1.12) events was observed following vaccination. The risk of cardiovascular events (myocarditis, RR, 8.53; myocardial infarction, RR, 2.59; cardiac arrhythmia, RR, 4.47) after SARS-CoV-2 infection was much higher than after vaccination.

**Conclusion:** Although there is a risk of cardiovascular events following vaccination, the risk was much lower than that following SARS-CoV-2 infection. The benefits of COVID-19 vaccination to the population outweigh the risks in terms of cardiovascular safety assessment.

## 1 Introduction

WHO reported that, as of 23 June 2022, there have been a total of more than 539 million confirmed cases of COVID-19 including 6,324,112 cumulative deaths worldwide^[1]^. COVID-19 vaccination with an effective and well-tolerated vaccine is an effective measure to reduce the rate of infection in the population and to contain the coronavirus pandemic. Moreover, several large studies have evaluated that clinical trials of the COVID-19 vaccine have demonstrated a good safety^[2]^ and efficiency^[3]^, and that full vaccination is effective in preventing the incidence of severe illness and mortality after COVID-19 infection.

However, as cardiovascular events after vaccination have been reported^[2,4–7]^. The potential adverse event of vaccines raised hesitancy to vaccinate, especially in the elderly population. These may affect vaccination rates and may lead to a longer duration of the COVID-19 pandemic. Moreover, COVID-19 infection increases the risk of cardiovascular events. In addition, the recent emergence of more infectious and immune evasive mutants has made the management of outbreaks more difficult. Therefore, a realistic and objective assessment of vaccine safety has become a major concern for the international community.

Randomized controlled trials are usually conducted under harsh and ideal conditions, with a high degree of consistency in population selection and study setting, which has some limitations in assessing vaccine safety. However, in the case of larger vaccinations in a population, the factors that need to be considered (e.g., individual willingness to vaccinate, group heterogeneity) are more complex than in RCTs. Real-world studies provide a better indication of the cardiovascular safety of the vaccine in vaccinated populations. Several studies have recently reported real-world cardiovascular safety assessments of the COVID-19 vaccine, but the results remain controversial^[8–11]^. Studies by Barda N and Karlstad Ø concluded that vaccination was associated with an elevated risk of myocarditis. However, lp S^[12]^ found little evidence to suggest higher incidence of these events after second vaccination. Therefore, meta-analyses of real-world studies remain necessary. In this study, we assessed the risk of MACE of COVID-19 vaccine to establish a reliable evidence base for the cardiovascular safety of COVID-19 vaccine in the real world.

## 2 Methods

### 2.1 Search strategy

This study was registered in the PROSPERO (CRD 42022344375). A systematic search was done according to the Preferred Reporting Items for Systematic Review and Meta-Analysis (PRISMA) statement^[13]^ for the conduct of meta-analyses of real-world studies. Searches were conducted of PubMed, EMBASE, Web of Science database, without language restriction from January 1, 2020 to June 15, 2022. Gray literature from Google Scholar, and preprint reports was added to the searches of electronic databases. The terms of Medical Subject Headings and relevant text words is consisted as following: “COVID-19 vaccination/vaccine”, “SARS-CoV-2 vaccination/vaccine”, “myocarditis, myopericarditis, cardiac inflammations”, “myocardial infarction”, “arrhythmia, cardiac arrhythmia” “cardiovascular events”.

### 2.2 Inclusion criteria and exclusion criteria

All clinical studies were included in the systematic review, while real-world studies were included in the meta-analysis. Intervention studies (such as randomized controlled studies), animal studies, comments, review articles, case reports, or studies without available data were excluded from the meta-analysis.

### 2.3 Data extraction and quality evaluation

For each included study, data extraction and quality evaluation were performed independently by two authors (YF.C and EW.H). Any disagreements were settled by consultation. Data sought were the first author, year of publication, country/region, type of study, study period, data sources, vaccine type, outcomes, observation window (days), dose number, vaccine type, population characteristics, sample size, events. Intra-study risk of bias was evaluated using the Joanna Briggs Institute (JBI) checklist^[14,15]^ for prevalence studies and the Newcastle-Ottawa Scale^[16]^ for cohort studies.

### 2.4 Data analysis

The incidence rate and risk ratio (RR) were used to estimate effect sizes. The heterogeneity between effect values across studies was estimated by I^2^ statistic. Fixed-effects models were used if I^2^ ≤ 50%. Random-effects models were used if I^2^ >50%, representing significant heterogeneity. Random-effects meta-regression analyses were used to investigate the associations of population size and vaccine sample size with the observed incidence rate and RR. Sample sizes for both population and doses were divided into three groups of less than 1 million, 1-10 million, and greater than 10 million. Subgroup analyses were performed by sex, age, dose number, and vaccine type to assess the incidence rate and myocarditis. Evidence of publication bias were examined by funnel plot, Begg’s test. The robustness and reliability of the meta-analysis results were evaluated by sensitivity analyses. Statistical analyses were performed with Stata version 17.0. For all analyses, P value of less than 0.05 was deemed significant.

## 3 Results

### 3.1 Search results and characteristics of included studies

The literature search screened 1296 studies of which 102 were reviewed in full text (Figure 1). A total of 43 studies were included in the meta-analyses, of which reported 16,978 cases of myocarditis^[7,8,17–51]^, 28,451 cases of myocardial infarction^[8,19,20,43,47,50,52–58]^ and 96,269 cases of cardiac arrhythmia^[8,19,21,22,24,43,44]^ (Table S1). Overall risk of bias in these studies were rated as moderate to high quality (Tables S2 and S3). The association between cardiovascular events and COVID-19 vaccination was evaluated in people aged 12 years or order. The population size ranged from 8553 to 557,868,504. Funnel plots and Begg’s test identified no strong evidence of publication bias. *P* values are all >0.05 (Figure S1). Sensitivity analyses showed no significant change in the results of the incidence and risk ratio of cardiovascular events after excluding each study one by one, indicating the robustness of the meta-analysis (Figure S2).

**Figure 1.**
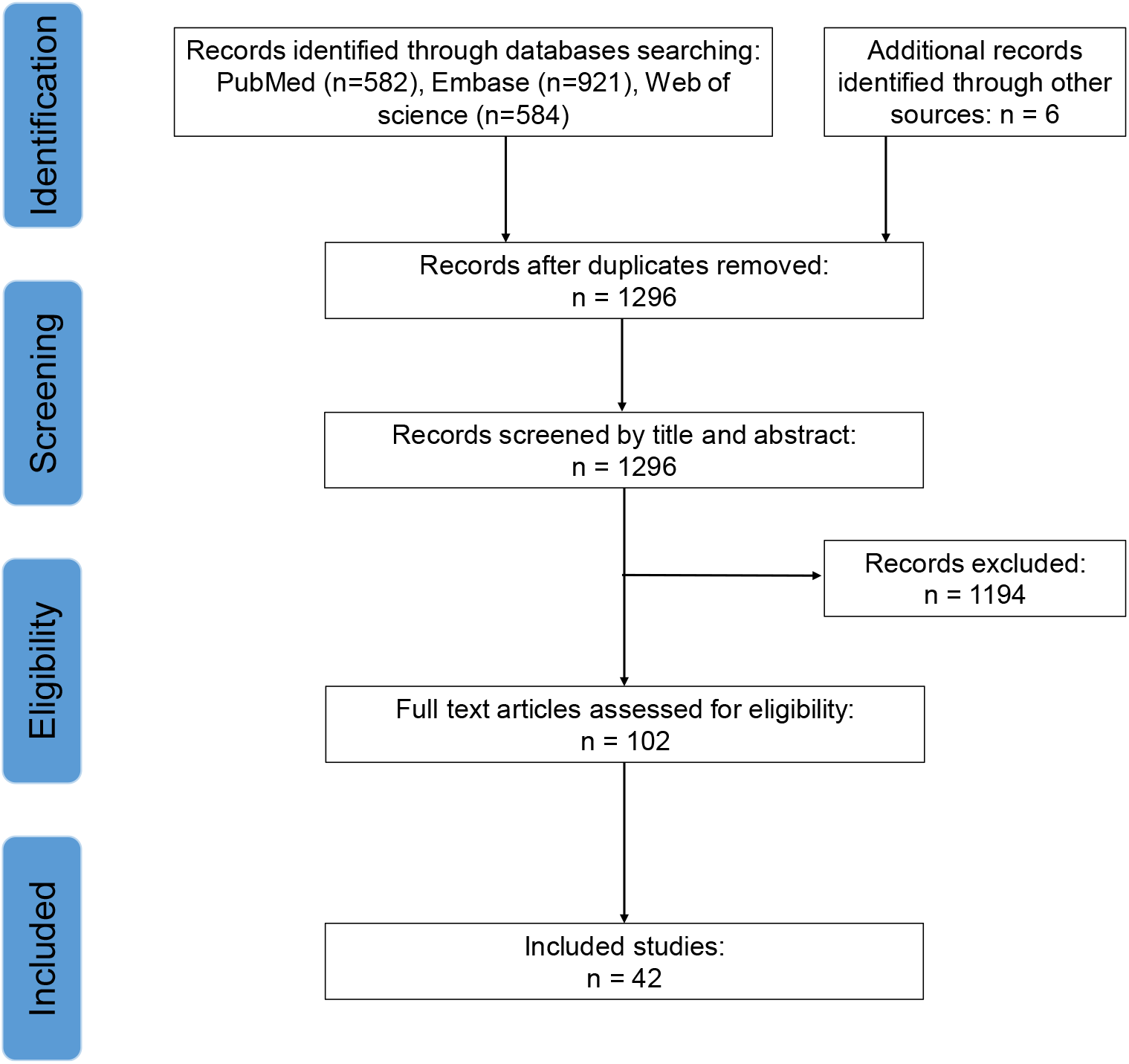
Article Identification flow chart following the PRISMA guidelines.

### 3.2 Association of myocarditis and COVID-19 vaccine

A total of 1,604,254,833 persons received 2,575,129,450 doses vaccine were eligible for inclusion in the studies. Of those with myocarditis, the median time to symptom onset was 3.2(0.6-5.8) days. The overall incidence rate of myocarditis after COVID-19 vaccination were 14.8(13.0-16.6) events per million persons, 8.8(7.8-9.9) events per million doses. The meta-regression analyses identified no association between population size (*P* = 0.057) or vaccine sample size (*P* = 0.277) and myocarditis incidence rate. Subgroup analyses indicated that the incidence of myocarditis was more frequently in males than in females (18.1(14.5-21.8) vs. 4.9(3.7-6.0) events per million people, *P*<0.001), and in aged 12-39 years than in 40 years or older (17.7(11.1-24.4) vs. 5.2(1.5-8.9) events per million people, *P*<0.001). During the risk interval, there were 16,978 cases of myocarditis corresponding to incidence rate of 5.5 events per million doses after first vaccination, 13.7 events per million doses after second vaccination, and 5.9 events per million doses after third vaccination (Figure 2). Two of the included studies reported the events rate (107.9(61.8-154.0) events per million persons) of myocarditis following SARS-CoV-2 infection.

**Figure 2.**
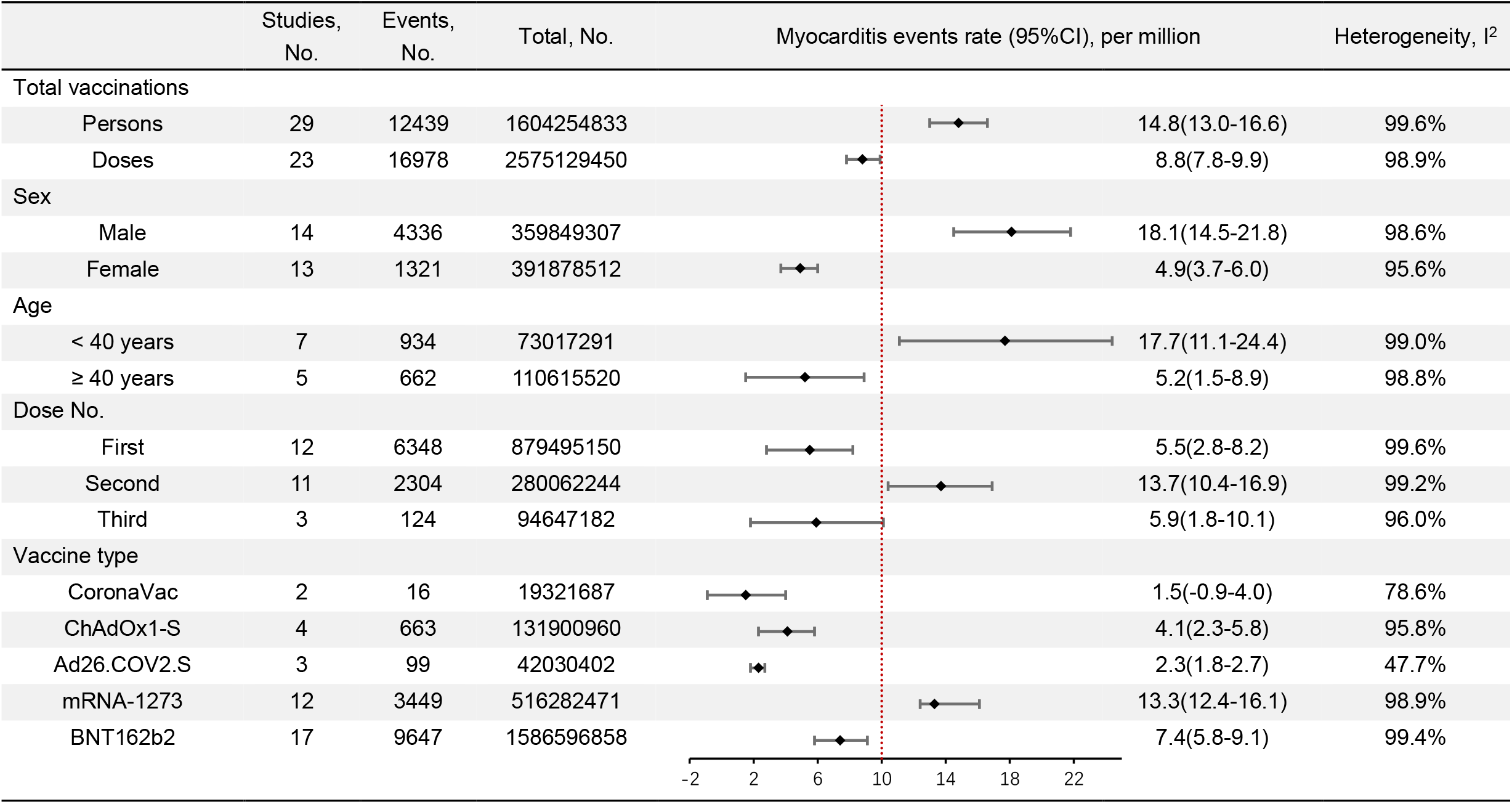
Incidence rate for myocarditis events after COVID-19 vaccination.

Among the included studies, 17 studies on vaccination and the myocarditis risk. Figure 3 shows the risk ratios in the vaccination analyses for myocarditis events. In the total population, the risk of myocarditis events was higher in the vaccinated than in the unvaccinated, with a risk ratio of 1.39(95% CI, 1.13 -1.65). The meta-regression analyses identified no association between population size and myocarditis risk ratio (*P* = 0.082). Subgroup analyses by dose and vaccine type found higher risks of myocarditis following second vaccination (RR, 2.09; 95%CI: 1.59-2.58), third vaccination (RR, 2.02; 95%CI: 1.40-2.91) compared without vaccination. Furthermore, there were increased risks of myocarditis following a second dose of mRNA-1273 (RR, 7.27; 95%CI:5.33-9.21), of BNT162b2 (RR, 1.55; 95%CI:1.36-1.74) (Table S4). There were no evidence for myocarditis risk following CoronaVac or ChAdOx1-S vaccination. The increased risk myocarditis events following vaccination were higher in males than in females (RR, 3.44; 95%CI: 2.61 to 4.54) and in younger populations (<40 vs. ≥40 years: RR, 2.20; 95%CI: 1.06-4.55). Among the unvaccinated, the risk ratio was higher in males (RR, 1.71; 95%CI: 1.49-1.93), while lower in younger (<40 vs. ≥40 years: RR, 0.72; 95%CI: 0.61-0.83). Four of the included studies reported the risk of myocarditis following SARS-CoV-2 infection. The combination revealed that infection substantially increased the risk of myocarditis (RR, 8.53; 95%CI: 2.15-14.91).

**Figure 3.**
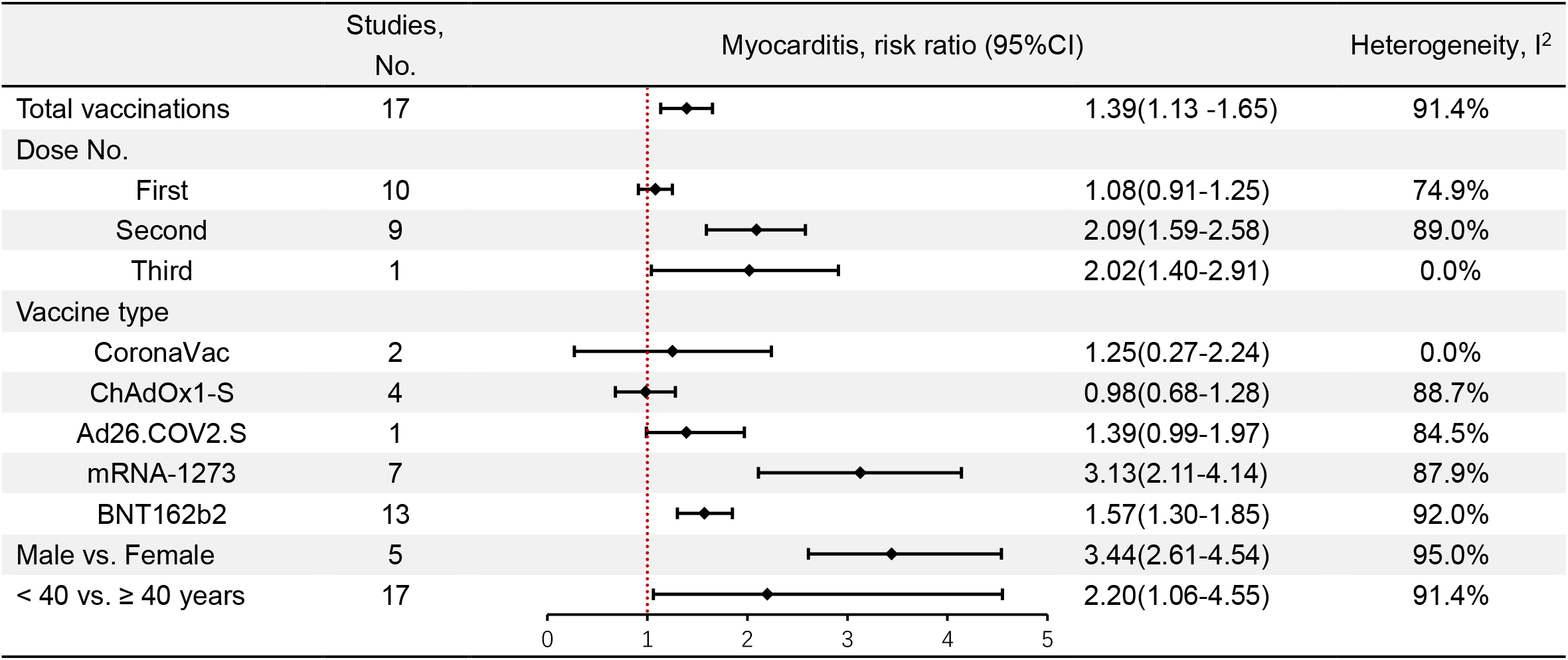
Risk ratio for myocarditis events after COVID-19 vaccination.

### 3.3 Association of myocardial infarction and COVID-19 vaccine

A total of 610,438,891 persons received 670,082,343 doses vaccine were included in the myocardial infarction events analyses. The overall incidence rate of myocardial infarction after COVID-19 vaccination were 1.73(1.63-1.82) events per 10,000 persons, 1.62(1.46-1.77) events per 10,000 doses. The meta-regression analyses identified no association between population size (*P* = 0.643) or vaccine sample size (*P* = 0.822) and myocardial infarction incidence rate. Subgroup analyses by dose found myocardial infarction incidence rate after first vaccination and second vaccination were 1.81 and 1.10 events per million doses, respectively (Table 1). The incidence rate associated with SARS-CoV-2 infection (9.81 events per 10,000 persons) were much higher than that associated with the vaccine (*P* < 0.001).

**Table 1.** Incidence rate for myocardial infarction events following COVID–19 vaccinations.

|  | Studies,<br>No. | Events,<br>No. | Total, No. | Events rate (95%CI), per 10,000 | Heterogeneity, I<sup>2</sup> |
| --- | --- | --- | --- | --- | --- |
| Total vaccinations |  |  |  |  |  |
| Persons | 11 | 26698 | 610438891 | 1.73(1.63–1.82) | 100% |
| Doses | 10 | 28451 | 670082343 | 1.62(1.46–1.77) | 100% |
| Sex |  |  |  |  |  |
| Male | 2 | 1160 | 10371966 | 1.03(0.78–1.28) | 71.8% |
| Female | 2 | 394 | 10722873 | 0.37(0.33–0.40) | 71.8% |
| Age |  |  |  |  |  |
| < 40 years | 2 | 100 | 10792995 | 0.09(0.07–0.11) | 0.0% |
| ≥ 40 years | 5 | 2733 | 21573293 | 1.54(0.09–2.98) | 99.4% |
| Dose No. |  |  |  |  |  |
| First | 5 | 9160 | 45715167 | 1.81(0.06–3.55) | 100% |
| Second | 3 | 1244 | 18207826 | 1.10(0.02–2.22) | 99.3% |
| Vaccine type |  |  |  |  |  |
| CoronaVac | 1 | 8764 | 70423565 | 0.38(0.35–0.41) | 100% |
| ChAdOx1-S | 3 | 7956 | 22352515 | 2.12(-1.64–5.87) | 100% |
| Ad26.COV2.S | 1 | 128 | 5891211 | 0.22(0.18–0.26) | 0.0% |
| mRNA-1273 | 2 | 548 | 131842583 | 0.98(0.91–2.87) | 97.4% |
| BNT162b2 | 7 | 10275 | 647096265 | 0.96(0.90–1.03) | 99.9% |

No association was observed with the vaccine in total population (RR, 0.98; 95%CI: 0.87-1.09), including first vaccination (RR, 0.97; 95%CI: 0.86-1.09) and second vaccination (RR, 1.06; 95%CI: 0.99-1.14). No association was found with the ChAdOx1-S (RR, 0.91; 95%CI: 0.76-1.06) or BNT162b2 vaccines (RR, 0.95; 95%CI: 0.85-1.05). The risk ratio of myocarditis events following vaccination were higher in males (RR, 2.61; 95%CI: 1.65-4.14) and in persons aged over 40 years (≥40 vs. <40 years: RR, 11.73; 95%CI: 4.83-28.48). Among the unvaccinated, the risk was higher in males than in females (RR, 2.86; 95%CI: 1.54-4.18) and in those older than 40 (≥40 vs. <40 years: RR, 5.23; 95%CI: 4.98-5.49) for MI (Figure 4). Two of the included studies reported the risk of myocardial infarction following SARS-CoV-2 infection. The combination revealed that infection was associated with a substantially increased risk of myocardial infarction (RR, 2.59; 95%CI: 2.38-2.79).

**Figure 4.**
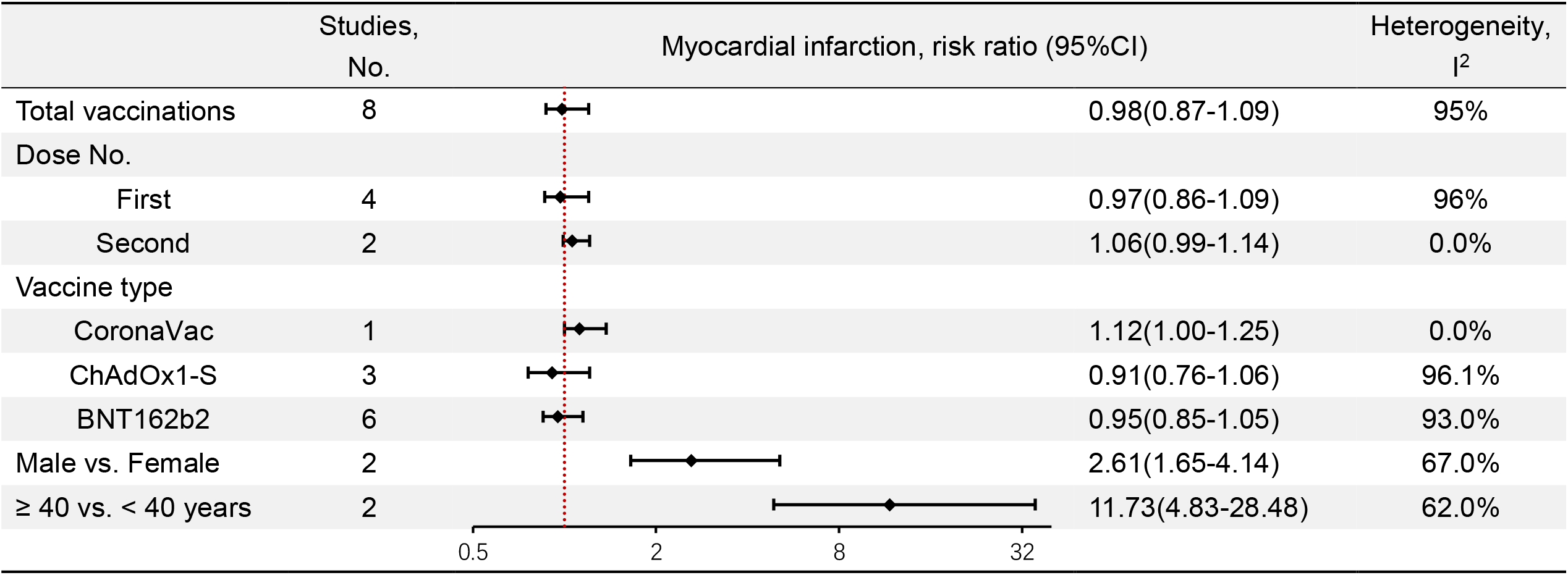
Risk ratio for myocardial infarction events following COVID-19 vaccination.

### 3.4 Association of cardiac arrhythmia and COVID-19 vaccine

A total of 59,951,135 persons and 372,839,175 doses vaccine were included in the cardiac arrhythmia study. The overall incidence rate of cardiac arrhythmia after COVID-19 vaccination were 9.62 events per 10,000 persons and 5.09 events per 10,000 doses. Subgroup analyses by vaccination dose found cardiac arrhythmia incidence rate after first vaccination and second vaccination were 6.07 and 5.04 events per million doses, respectively (Table 2).

**Table 2.** Incidence rate for cardiac arrhythmia events following COVID–19 vaccinations.

|  | Studies,<br>No. | Events,<br>No. | Total, No. | Events rate (95%CI),<br>per 10,000 | Heterogeneity, I<sup>2</sup> |
| --- | --- | --- | --- | --- | --- |
| Total vaccinations |  |  |  |  |  |
| Persons | 4 | 88726 | 59951135 | 9.6(-4.3–23.6) | 100% |
| Doses | 5 | 96269 | 372839175 | 5.1(3.2–7) | 100% |
| Sex |  |  |  |  |  |
| Male | 3 | 46401 | 27886355 | 9.9(-8.4–28.1) | 100% |
| Female | 3 | 41965 | 31787342 | 7.8(-6.5–22.1) | 100% |
| Age |  |  |  |  |  |
| < 40 years | 3 | 5009 | 22452974 | 2.1(-0.9–5.2) | 100% |
| ≥ 40 years | 3 | 83383 | 44570456 | 11.7(-10.9–34.3) | 100% |
| Dose No. |  |  |  |  |  |
| First | 4 | 43848 | 59193521 | 6.1(-1.1–13.2) | 100% |
| Second | 3 | 44643 | 47214867 | 5(-5.3–15.4) | 100% |
| Vaccine type |  |  |  |  |  |
| CoronaVac | 1 | 485 | 17030243 | 0.3(0.3–0.3) | 0% |
| ChAdOx1-S | 2 | 50215 | 91159837 | 4.2(-0.1–8.4) | 100% |
| Ad26.COV2.S | 1 | 144 | 5891211 | 0.2(0.2–0.3) | 0% |
| mRNA-1273 | 2 | 375 | 1513526 | 6.9(-3.8–17.5) | 99.2% |
| BNT162b2 | 5 | 45047 | 257165581 | 5.3(3.1–7.6) | 100% |

No association was observed of cardiac arrhythmia and the vaccine in total population (RR, 0.98; 95%CI: 0.84-1.12), including first vaccination (RR, 0.97; 95%CI: 0.91-1.02) and second vaccination (RR, 0.97; 95%CI: 0.94-1.01). No association was found with the ChAdOx1-S, mRNA-1273 or BNT162b2 vaccines. Among the vaccinated population, the risk ratio of cardiac arrhythmia events was higher in older individuals (≥40 years vs.<40: RR, 3.83; 95%CI: 1. 82-8.07), similar results in unvaccinated individuals (≥40 years vs.<40: RR, 2.27; 95%CI: 1.78-2.89) (Figure 5). Three of the included studies reported the risk of cardiac arrhythmia following SARS-CoV-2 infection. The combination revealed that infection substantially increased the risk of cardiac arrhythmia (RR, 4.47; 95%CI: 3.32-5.63).

**Figure 5.**
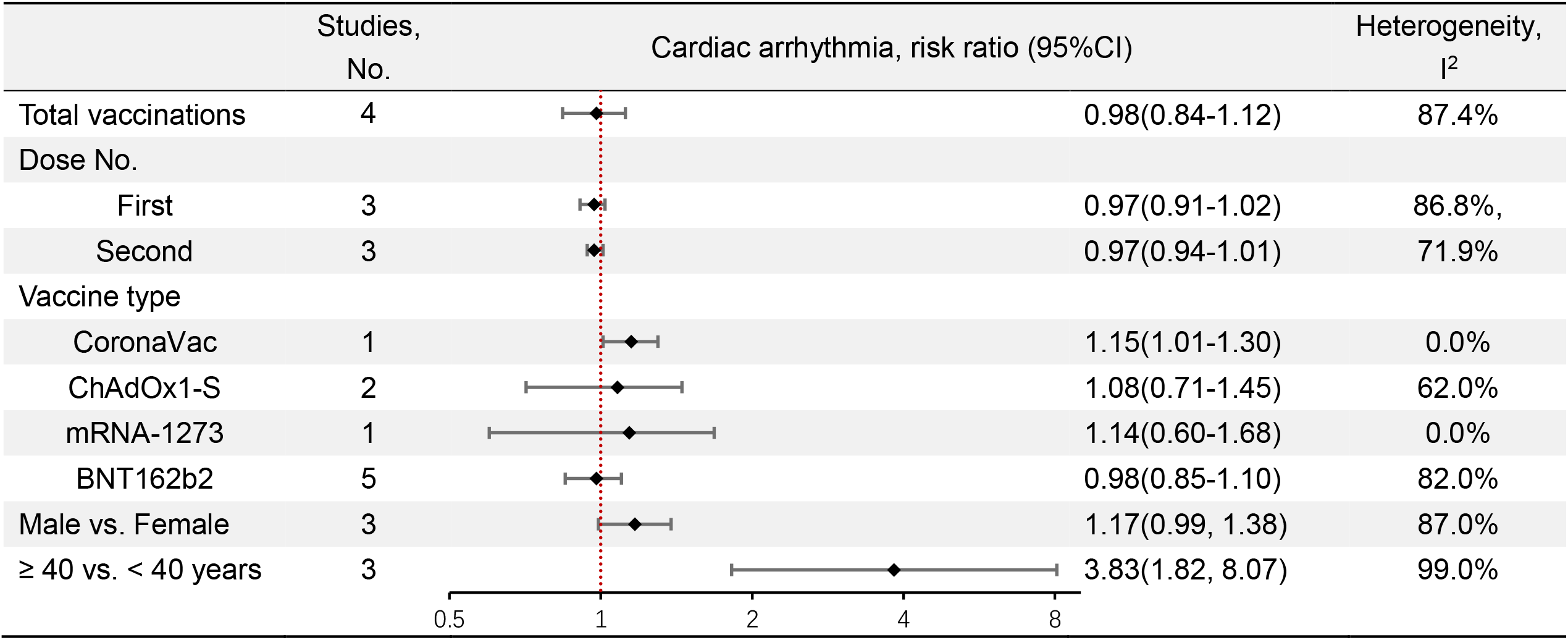
Risk ratio for cardiac arrhythmia events following COVID-19 vaccination.

## 4 Discussion

In this systematic review and meta-analysis, we examined the association between COVID-19 vaccines and MACE to evaluate the cardiovascular safety profile of COVID-19 vaccine. The study results showing an increased risk of myocarditis after vaccination, but it is a rare adverse event with a rate of 14.8 events per million persons. In the general population, the increased risk of myocarditis after vaccination was mostly in young men and observed following the second and third dose. Our results did not indicate statistically significant associations between vaccine and MI or cardiac arrhythmias events. The incidence and risk of COVID-19-associated MACE were greatly higher than that of COVID-19 vaccine-related MACE.

Vaccine-induced myocarditis is suspected to be the result of an autoimmune phenomenon. Rare cases of myocarditis have also been reported after vaccination for other diseases. However, verification of a causal relationship between myocarditis and the COVID-19 vaccine is not possible. The Potential mechanisms might be mRNA immune reactivity, antibodies to SARS-CoV-2 spike glycoproteins cross-reacting with myocardial contractile proteins, and hormonal differences^[59]^. In addition, Can et al^[60]^ suggests that COVID-19 mRNA vaccine-associated myopericarditis in murine model may be due to inadvertent intravenous administration or rapid return from the lymphatic circulation resulting in elevated systemic levels of mRNA lipid nanoparticles.

The findings from the present systematic review show that, the CoronaVac vaccine had the lowest incidence of myocarditis, while the Ad26.COV2.S vaccine had the lowest incidence and risk of myocardial infarction and cardiac arrhythmias. Multiple studies have shown that, inactivated vaccines had the highest safety profile and the lowest incidence of myocarditis and myocardial infarction events ^[61–65]^. The safety comparisons between mRNA vaccines and adenovirus vector vaccines were controversial. A head-to-head comparison^[34]^ of BNT162b2 and mRNA-1273 vaccines indicated that myocarditis risk was higher after mRNA-1273 vaccination than after BNT162b2 vaccination. Both vaccines were associated with an excess risk of myocarditis^[2]^. While, another head-to-head comparison^[66]^ for men aged 18 to 25 years do not indicate a statistically significant difference in myocarditis risk between recipients of mRNA-1273 and BNT162b2. This study demonstrated an increased risk of myocarditis after mRNA-1273 vaccination than after BNT162b2 vaccination. Possibly due to the heterogeneity of the population: Individuals receiving ChAdOx1 or BNT162b2 vaccines are on average older than those receiving mRNA-1273 vaccine; The interval between the first and second dose is different in BNT162b2 vaccine (3 weeks) and mRNA-1273 (4 weeks). In our meta-analysis, the incidence of myocarditis was 14.8 events per million persons following at least one dose vaccination. Data from a Large Health Care Organization of Israeli^[67]^ indicated the estimated incidence was 2.13 events per 100,000 persons. These rates are much lower than the incidence rate described for SARS-CoV-2-infected myocarditis.

Our analysis concluded that time from last vaccination to onset of myocarditis symptoms was 3.2 days. Most cases were considered mild or moderate in severity, with a short duration^[67,68]^. A systematic evaluation by Pillay^[69]^ concluded that most myocarditis occurs within the first week after vaccination, usually on day 3-4.The overall survival rate of patients with COVID-19 mRNA vaccine-associated myocarditis was greater than 99% ^[59]^. Rosenblum^[70]^ found that from VAERS reports, more than half of the patients showed mild adverse reactions and a short duration of symptoms.

Among the vaccinated population, the incidence of MACE varies greatly depending on age and sex. These events all occurs more commonly in males compared with females. The increased risk of myocarditis after vaccination was higher in persons aged under 40 years. The results are consistent with the characteristics of myocarditis in most studies^[44,71]^, while the explanation is still unclear. A few studies have concluded that the mechanism might the inhibition effects of testosterone on anti-inflammatory cells and the immune response to Th1 type cells in males, and the inhibitory effects of estrogen on proinflammatory T cells in females^[72]^.

We found no evidence of an increased risk of any MACE after the first administration. People who received the vaccine had the highest incidence and risk ratio for myocarditis after the second vaccination, followed by the third. Most studies^[23,66,73]^ have shown a higher risk of myocarditis after the second dose of mRNA vaccine. Such as, a cohort study from the United States^[66]^ concluded that among young men, the pooled incidence rate was highest after the second dose for BNT162b2 and mRNA-1273. However, only a few studies^[43,44]^ reported statistically insignificant risk for myocarditis and the second dose vaccine, especially CoronaVac and ChAdOx1-S vaccine. Most of the studies included in our review did not report on cardiovascular outcomes after receiving the third dose. Therefore, further studies are needed to determine the outcomes of the third dose. In this study, no significant increased risk of myocardial infarction or arrhythmia was found after any vaccination. There was a greater risk of myocarditis (RR, 8.53), myocardial infarction (RR, 2.59), cardiac arrhythmia (RR, 4.47) following SARS-CoV-2 infection, and that virus significantly increases the risk of many other serious adverse events^[2]^. Even among the young male population, COVID-19-associated myocarditis were 6 times higher than that of COVID-19 vaccine-related myocarditis^[74]^.

We conducted random-effects models for meta-analysis and implemented multiple subgroup analyses to assess potential sources of heterogeneity. The heterogeneity might be due to the diversity of study populations, follow-up, vaccine component, diagnostic criteria. Comparisons between incidence rates from different sources may yield systematic errors. In addition, some of the studied outcomes also had population-level heterogeneity that followed age patterns. The incidence of myocardial infarction and cardiac arrhythmia increased with age. In this analysis, the heterogeneity did not change significantly after our subgroup analysis by sex, age, and vaccine type. However, the limited number of studies in some subgroups limited the power of such subgroup analysis.

Limitations of this study. First, studies on cardiovascular events of the vaccine were included in this meta-analysis, while studies on other side effects of the vaccine were not included. Second, the significant heterogeneity observed across studies could not explored nor explained by subgroup analyses or meta-regression. Third, most studies did not report outcomes in people under 12 years of age who were vaccinated.

In conclusion, the present study suggests that myocarditis following COVID-19 vaccination is rare. The highest myocarditis risk was in men aged under 40 years after second dose. No significant increase for the three cardiovascular events was observed following first vaccination. No significant increased risk of myocardial infarction or arrhythmia was found after any vaccination. The risk of cardiovascular events was much higher after SARS-CoV-2 infection than after COVID-19 vaccination. The benefits of COVID-19 vaccination to the population outweigh the risks in terms of cardiovascular safety assessment.

## Data Availability

All data produced in the present study are available upon reasonable request to the authors

## Acknowledgments

This study was supported by the Natural Science Foundation of Guangdong Province (Grant No. 2018A030313254) and the Guangzhou Science and Technology Program (Grant No. 2019030015).

