## Supplementary material for "cardiovascular safety of COVID-19 vaccines in real-world studies: a systematic review and meta-analysis": Table S 1-4, Figure S 1-2

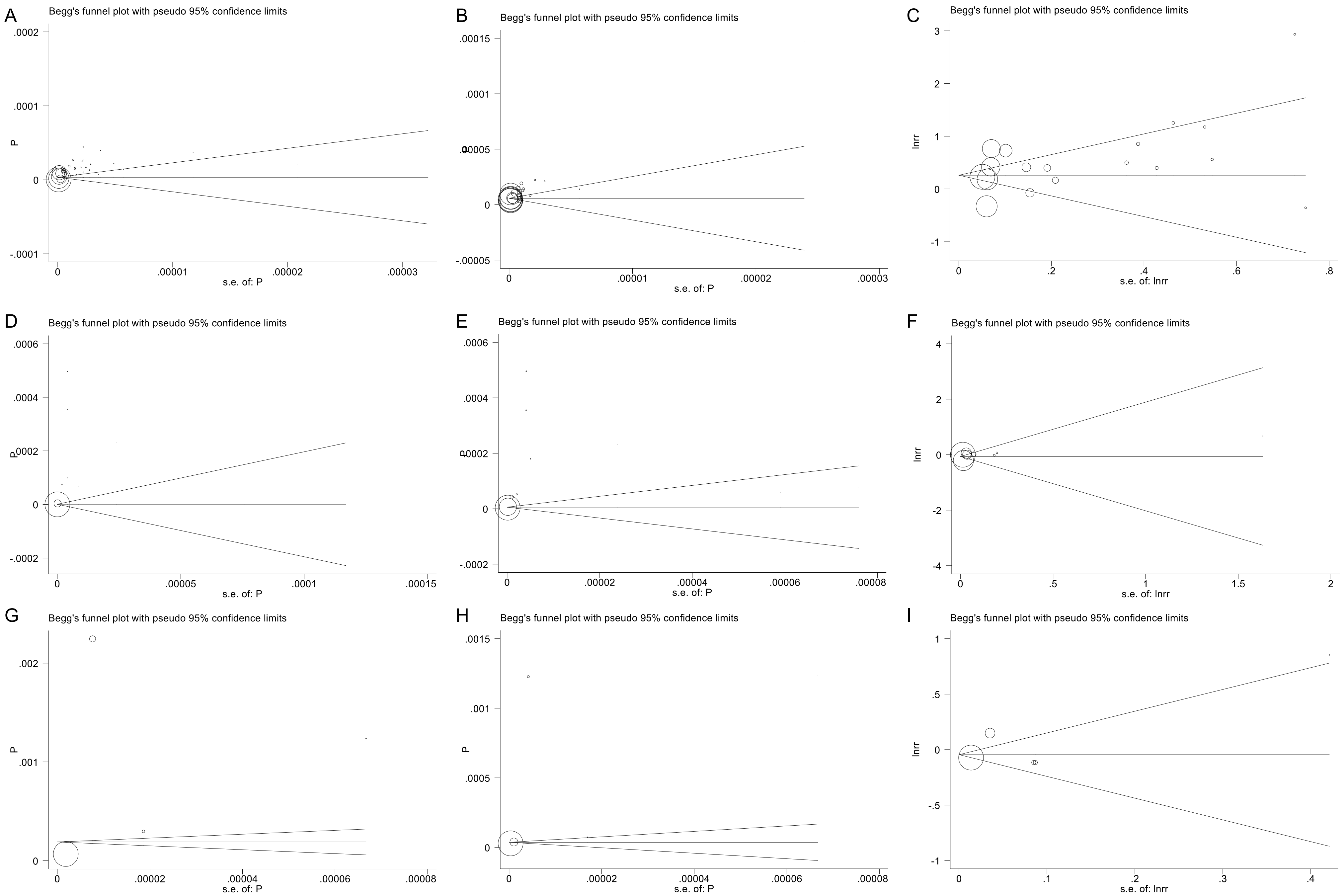

Figure S1. Funnel plot and Begg's test for (A) myocarditis incidence rate in total persons (*P* = 0.063); (B) myocarditis incidence rate in total doses (*P* = 0.102); (C) myocarditis risk ratio (*P* = 0.544); (D) myocardial infarction incidence rate in total persons (*P* = 0.533); (E) myocardial infarction incidence rate in total doses (*P* = 1.000); (F) myocardial infarction risk ratio (*P* = 0.174); (G) cardiac arrhythmia incidence rate in total persons (*P* = 0.734); (H) cardiac arrhythmia incidence rate in total doses (*P* = 0.221); (I) cardiac arrhythmia risk ratio (*P* = 0.462).

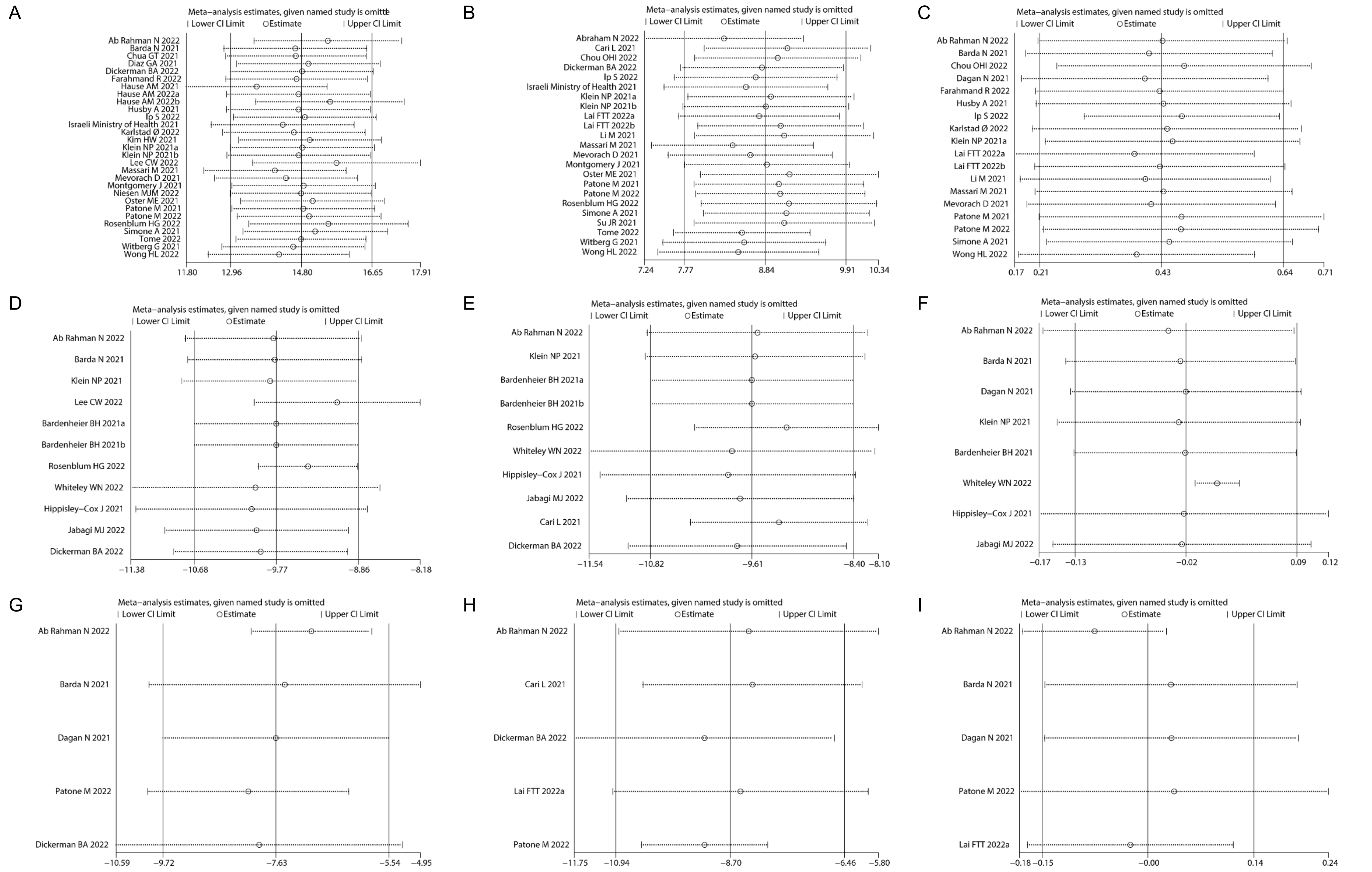

Figure S2. Sensitivity analysis for (A) myocarditis events rate in total persons; (B) myocarditis events rate in total doses; (C) myocarditis risk ratio; (D) myocardial infarction events rate in total persons; (E) myocardial infarction events rate in total doses; (F) myocardial infarction risk ratio; (G) cardiac arrhythmia incidence rate in total persons; (H) cardiac arrhythmia events rate in total doses; (I) cardiac arrhythmia risk ratio.

**Table S1. Characteristics of included studies.**

| Author | Study period | Country/ Region | Data source | Study design | Outcomes; Observation window (days) | Population, age | Dose No. | Vaccine type | Total (N) | Myocarditis events (N) | Myocardial infarction events (N) | Cardiac arrhythmia events (N) |
|---|---|---|---|---|---|---|---|---|---|---|---|---|
| Ab Rahman N 2022 | February 1, 2021 to September 30, 2021 | Malaysia | the Malaysian Data Warehouse | SCCS | Myocardial infarction, Myocarditis / pericarditis, Arrhythmia; 21 days following either vaccination | ≥12 years | first, second | BNT162b2, CoronaVac, ChAdOx1 | 20202054 people, 35201509 doses | 25 | 1495 | 1375 |
| Abraham N 2022 | December 2020 to March 13, 2022 | Canada | Canadian Adverse Events Following Immunization Surveillance System | observational study | Myocarditis / pericarditis; 7 days post-vaccination | 18-39 years | first, second | BNT162b2, mRNA-1273 | 19370047 | 372 | | |
| Barda N 2021 | December 20, 2020 to May 24, 2021 | Israel | Clalit Health Services in Israel | observational cohort study | each potential adverse event; 42 days after vaccination | ≥16 years | first, second | BNT162b2 | 938812 persons | 21 | 59 | 254 |
| Bardenheier BH 2021a | December 18, 2020 to March 7, 2021 | US | Electronic Health Record data from Genesis HealthCare | cohort study | mortality of 7 days post-vaccination, Other adverse events of 15 days post-vaccination | average age ≥60 years | first, second | Moderna, Pfizer-BioNTech | 8553 persons, 16924 doses | | 1 | |
| Bardenheier BH 2021b | December 18, 2020 to February 14, 2021 | US | a large nursing home provider spanning 24 US states | retrospective cohort study | mortality of 7 days post-vaccination, Other adverse events of 15 days post-vaccination | average age ≥60 years | first | Moderna, Pfizer-BioNTech | 13163 persons | | 1 | |
| Cari L 2021 | to June 21, 2021 | European | EudraVigilance, the European Centre for Disease Prevention and | observational study | Adverse events, including myocardial infarction and cardiac arrhythmia | ≥18years | at least one dose | ChAdOx1, Ad26.COV2.S, BNT162b2 | 266391184 dose | | 1871 | 7778 |

| | | | | | | | | | | | | |
|---|---|---|---|---|---|---|---|---|---|---|---|---|
| | | | Control database | | | | | | | | | |
| Chou OHI 2022 | January 1, 2020 to June 30, 2021 | Hong Kong, China | any of the Hong Kong public hospitals, local electronic healthcare database | retrospective cohort study | Myopericarditis; time interval ≤14 days | | first, second | CoronaVac, BNT162b2 | 7588200 doses | 42 | | |
| Chua GT 2021 | June 14, 2021 to September 4, 2021 | Hong Kong, China | Comirnaty vaccination | observational study | myocarditis/pericarditis; ≤14 days | 12-17 years | first, second | BNT162b2 | 178163 persons, 305406 doses | 33 | | |
| Dagan N 2021 | December 20, 2020 to May 24, 2021 | Israel | Clalit Health Services in Israel | observational cohort study | each potential adverse event; 42 days after vaccination | ≥16 years | first, second | BNT162b2 | 938812 persons | 21 | 59 | 254 |
| Diaz GA 2021 | February 2021 to May 2021 | US | Forty hospitals of American | observational study | myocarditis, myopericarditis, pericarditis | IQR 57(40-70) years | first, second | Ad26.COV2.S, mRNA-1273, BNT162b2 | 2000287 persons | 20 | | |
| Dickerman BA 2022 | January 4, 2021 to September 20, 2021 | US | the national health care databases of the US Department of Veterans Affairs | observational study | myocardial infarction, other thromboembolic events, myocarditis or pericarditis, arrhythmia, other adverse events; 14 days after receipt of the first vaccine dose | ≥18 years | first | BNT162b2, mRNA-1273 | 429564 | 6 | 94 | 343 |
| Farahmand R 2022 | August 3, 2020 to May 21, 2021 | Israel | Beth Israel Deaconess Medical Center; Massachusetts Immunization Information System | cohort study | Incidence of Myopericarditis and Myocardial Injury in Coronavirus Disease 2019 Vaccinated Subjects | ≥18 years | first, second | BNT162b2, mRNA-1273, ChAdOx1, Ad26.COV2.S | 268320 persons | 10 | | |

| | | | | | | | | | | | |
|---|---|---|---|---|---|---|---|---|---|---|---|
| Hause AM 2021 | December 14, 2020 to July 16, 2021 | US | VAERS | observational study | myocarditis | 12-17 years | first, second | BNT162b2 | 8900000 persons | 397 | |
| Hause AM 2022a | December 9, 2021 to February 20, 2022 | US | VAERS | observational study | myocarditis | 12-17 years | booster | BNT162b2 | 2800000 persons | 47 | |
| Hause AM 2022b | September 22, 2021 to February 6 | US | VAERS | observational study | myocarditis | ≥18 years | booster | BNT162b2, mRNA-1273 | 81200000 persons | 37 | |
| Hippisley-Cox J 2021 | December 20, 2020 to May 24, 2021 | UK | the Office for National Statistics, the United Kingdom's health service | SCCS | adverse events; 21 days of follow-up after each of the first and second vaccine doses | ≥16years | first | ChAdOx1 n、BNT162b2 | 29121633 persons | | 14445 |
| Husby A 2021 | October 1, 2020 to October 5, 2021 | Denmark | Danish healthcare system | cohort study | The primary outcome, myocarditis or myopericarditis; follow-up time 0-28 days from the day of vaccination | ≥12 years | first, second | BNT162b2, mRNA-1273, ChAdOx1, Ad26.COV2.S | 4155361 persons | 69 | |
| Israeli Ministry of Health 2021 | December 2020 to May 2021 | Israel | the Ministry of Health database of Israel | observational study | myocarditis events，from first dose to 30 days after second doses | ≥16 years | first, second | mRNA vaccines | 5401150 persons | 148 | |
| Jabagi MJ 2022 | December 15, 2020 to April 30, 2021 | French | the French National Health Data System | SCCS | acute myocardial infarction, stroke, or pulmonary embolism; 14 days after vaccination | ≥75 years | first, second | BNT162b2 | 3900000 persons | | 1277 |
| Karlstad Ø 2022 | December 27, 2020 to October 5, 2021 | Denmark, Finland, | nationwide health registers | cohort study | myocarditis or pericarditis | ≥ 12 years | first, second | BNT162b2，mRNA- | 18814068 persons | 347 | |

| | | | | | | | | | | | | |
|---|---|---|---|---|---|---|---|---|---|---|---|---|
| | | Norway, Sweden | | | | | | 1273, AZD1222 | | | | |
| Kim HW 2021 | February 1, 2021 to April 30, 2021 | US | Duke University Medical Center | prospective study | myocarditis | ≥16 years | first, second | mRNA-1273, BNT162b2 | 561197 persons | 4 | | |
| Klein NP 2021a | December 14, 2020, to June 26, 2021 | US | eight data-contributing health plans of US | observational study | myocarditis/pericarditis; 42 days after first dose, 22 days after second dose | all ages, mean age 49 years | first, second | BNT162b2, mRNA-1273 | 6175813 persons, 11845128 doses | 87 | 613 | |
| Klein NP 2021b | December 2020 to August 21, 2021 | US | CDC | observational study | myocarditis/pericarditis, First in 60 Days | ≥12 years | first, second | Janssen, BNT162b2, mRNA-1273 | 7077839 persons, 13334831dose | 115 | | |
| Lai FTT 2022a | to September 30, 2021 | Hong Kong, China | Department of Health of the Hong Kong, Hospital Authority | retrospective cohort study | Adverse events, including myocarditis and cardiac arrhythmia | 12-18 years | first, second | BNT162b2 | 138141 dose1, 119664 dose2 | 38 | | 19 |
| Lai FTT 2022b | February 23, 2021 to August 2, 2021 | Hong Kong, China | Hospital Authority of Hong Kong | case-control study | carditis (acute myocarditis or pericarditis); | ≥12 | first, second | BNT162b2, CoronaVac | 9284702 | 27 | | |
| Lee CW 2022 | December 14, 2020 to September 30, 2021 | Korea | VAERS | observational study | Myocarditis/pericarditis; | ≥12 years | first, second | BNT162b2 | 235285732 persons | 474 | 59 | |
| Li M 2021 | December11, 2020 to August 13, 2021 | US | VAERS and CDC COVID Data Tracker | observational study | myocarditis and pericarditis | ≥12 years | first, second | Ad26.COV2.S, mRNA-1273, BNT162b2 | 353846154 doses | 2116 | | |
| lp S 2022 | December 8, 2020 to May 17, 2021 | England | Health Data Research UK and other institutions | cohort study | hospitalised or fatal myocarditis/pericarditis (0-13 days, 14+ days) | > 12 years | first, second | BNT162b2, ChAdOx1 | 49786346 persons | 607 | | |

| | | | | | | | | | | | | |
|---|---|---|---|---|---|---|---|---|---|---|---|---|
| | | | | | after first and second vaccinations | | | | | | | |
| Massari M 2021 | December 27, 2020 to September 30, 2021 | Italy | National Centre for Drug Research and Evaluation | SCCS | myocarditis/pericarditis; 0-21 days from the vaccination day | 12-39 years | first, second | BNT162b2, mRNA-1273 | 2861809 persons, 5109231 doses | 114 | | |
| Mevorach D 2021 | December 20, 2020, to May 31, 2021 | Israel | Ministry of Health database of Israel | retrospective study | myocarditis; 0-21 days after first dose and 0-30 days after second dose | ≥16 years | first, second | BNT162b2 | 5442696 persons | 136 | | |
| Montgomery J 2021 | January 2021 to April 2021 | US | US Military Health System, VAERS | retrospective study | myocarditis; | 20-51 years | first, second | BNT162b2, mRNA-1273 | 2810000 doses | 23 | | |
| Niesen MJM 2022 | December 2020 to October 2021 | US | multistate Mayo Clinic Enterprise | cohort study | adverse event within 14 days after each vaccine dose | IRQ 67 years | booster | BNT162b2, mRNA-1273 | 47999 persons | 1 | | |
| Oster ME 2021 | December 2020 to August 2021 | US | VAERS | observational study | myocarditis | ≥12 years | first, second | BNT162b2, mRNA-1273 | 192405448 persons, 354100845 doses | 1626 | | |
| Patone M 2021 | December 1, 2020 to November 15, 2021 | England | NIMS | SCCS | myocarditis, pericarditis, and cardiac arrhythmias, 1-28 days postvaccination period | ≥13 years | first, second, third | ChAdOx1, BNT162b2, mRNA-1273 | 42200614 persons; 91572102 doses | 552 | | |
| Patone M 2022 | December 1, 2020 to August 24, 2021 | England | NIMS | SCCS | myocarditis, pericarditis, and cardiac arrhythmias; 1-28 days postvaccination period | ≥16 years | first, second | ChAdOx1, BNT162b2, mRNA-1273 | 38615491 persons; 70711239 doses | 397 | | 86754 |
| Rosenblum HG 2022 | December 14, 2020 to June 14, 2021 | US | VAERS and v-safe | observational study | Adverse Event after 0-7day vaccination | ≥16 years | first, second | BNT162b2, mRNA-1273 | 298792852 doses | 1307 | 1118 | |

| | | | | | | | | | | | |
|---|---|---|---|---|---|---|---|---|---|---|---|
| Simone A 2021 | December 14, 2020 to July 20, 2021 | US | Kaiser Permanente Southern California | observational study | acute myocarditis; 0-10 days after vaccination | ≥18 years | first, second | BNT162b2, mRNA-1273 | 2392924 persons; 4629775 doses | 15 | |
| Su JR 2021 | December 2020 to October 6, 2021 | US | VAERS | observational study | myopericarditis, pericarditis; 0-7 days | ≥12 years | first, second | Janssen, BNT162b2, mRNA-1273 | 4024690 96 doses | 2459 | |
| Tome 2022 | January 1, 2021 to February 11, 2022 | EU/EEA countries | the EudraVigilance database and the European Centre for Disease Prevention and Control's vaccination tracker database | observational study | Myocarditis/pericarditis; | ≥12 years | first | mRNA-1273, BNT162b2 | 5578685 04 | 5356 | |
| Whiteley WN 2022 | December 8, 2020 to March 18, 2021 | England | General Practice Extraction Service Data for Pandemic Planning and Research | cohort study | major arterial, venous, and thrombocytopenic; 1 to 28 and >28 days after first vaccination | ≥18years | first | ChAdOx1-S、BNT162b2 | 2119381 4 persons | | 7536 |
| Witberg G 2021 | December 20, 2020 to May 24, 2021 | Israel | Clalit Health Services in Israel | retrospective cohort study | myocarditis; 21 days of follow-up after each of the two vaccine doses | ≥16 years | first, second | BNT162b2 | 2558421 persons | 54 | |
| Wong HL 2022 | Dec 14, 2021 to Jan 1, 2022 | US | four administrative claims databases (Optum, Health Core, Blue | retrospective cohort study | myocarditis, pericarditis; 1-7 days post-vaccination | 18-64 years | first, second | mRNA-1273, BNT162b2 | 1514836 9 persons, 2754427 0 doses | 411 | |

| | |
|---|---|
| | Health Intelligence, and CVS Health) |

SCCS: self-controlled case series study; VAERS: Vaccine Adverse Event Reporting System; CDC: Centers for Disease Control and Prevention; NIMS: National Incident Management System; EU: European Union; EEA: European Economic Area

Table S2. Joanna Briggs Institute Checklist for Prevalence Studies.

| Study | Was the sample frame appropriate to address the target population? | Were study participants sampled in an appropriate way? | Was the sample size adequate? | Were the study subjects and the setting described in detail | Was the data analysis conducted with sufficient coverage of the identified sample? | Were valid methods used for the identification of the condition? | Was the condition measured in a standard, reliable way for all participants? | Was there appropriate statistical analysis? | Was the response rate adequate, and if not, was the low response rate managed appropriately? | Overall |
|---|---|---|---|---|---|---|---|---|---|---|
| Ab Rahman N 2022 | √ | √ | √ | × | √ | √ | √ | √ | √ | 8 |
| Abraham N 2022 | √ | √ | √ | × | √ | √ | √ | √ | √ | 8 |
| Barda N 2021 | √ | √ | √ | √ | √ | √ | √ | √ | √ | 9 |
| Cari L 2021 | √ | √ | √ | √ | √ | √ | √ | √ | √ | 9 |
| Chua GT 2021 | √ | √ | √ | × | √ | √ | √ | √ | √ | 8 |
| Dagan N 2021 | √ | √ | √ | √ | √ | √ | √ | √ | √ | 9 |
| Diaz GA 2021 | √ | √ | √ | √ | √ | √ | √ | √ | √ | 9 |
| Dickerman BA 2022 | √ | √ | √ | √ | √ | √ | √ | √ | √ | 9 |
| Farahmand R 2022 | √ | √ | √ | √ | √ | √ | √ | √ | √ | 9 |
| Hause AM 2021 | √ | √ | √ | ? | ? | √ | √ | √ | √ | 7 |
| Hause AM 2022a | √ | √ | √ | ? | ? | √ | √ | √ | √ | 7 |
| Hause AM 2022b | √ | √ | √ | ? | ? | √ | √ | √ | √ | 7 |
| Husby A 2021 | √ | √ | √ | √ | √ | √ | √ | √ | √ | 9 |
| lp S 2022 | √ | √ | √ | √ | √ | √ | √ | √ | √ | 9 |
| Israeli Ministry of Health 2021 | √ | √ | √ | × | √ | ? | √ | √ | √ | 7 |
| Karlstad Ø 2022 | √ | √ | √ | √ | √ | √ | √ | √ | √ | 9 |
| Kim HW 2021 | √ | √ | √ | √ | √ | √ | √ | √ | √ | 9 |
| Klein NP 2021a | √ | √ | √ | √ | √ | √ | √ | √ | √ | 9 |
| Klein NP 2021b | √ | √ | √ | × | √ | √ | √ | √ | √ | 8 |
| Lai FTT 2022b | √ | √ | √ | | | | √ | √ | √ | |
| Lee CW 2022 | √ | √ | √ | × | √ | × | √ | √ | √ | 7 |
| Li M 2021 | √ | √ | √ | × | ? | × | √ | √ | √ | 6 |
| Massari M 2021 | √ | √ | √ | √ | √ | √ | √ | √ | √ | 9 |

| | | | | | | | | | | |
|---|---|---|---|---|---|---|---|---|---|---|
| Mevorach D 2021 | √ | √ | √ | ? | √ | √ | √ | √ | √ | 8 |
| Montgomery J 2021 | √ | √ | √ | √ | √ | √ | √ | √ | √ | 9 |
| Niesen MJM 2022 | √ | √ | √ | √ | √ | √ | √ | √ | √ | 9 |
| Oster ME 2021 | √ | √ | √ | √ | √ | √ | √ | √ | √ | 9 |
| Patone M 2021 | √ | √ | √ | × | √ | √ | √ | √ | √ | 8 |
| Patone M 2022 | √ | √ | √ | √ | √ | √ | √ | √ | √ | 9 |
| Rosenblum HG 2022 | √ | √ | √ | √ | √ | √ | √ | √ | √ | 9 |
| Simone A 2021 | √ | √ | √ | √ | √ | √ | √ | √ | √ | 9 |
| Su JR 2021 | √ | √ | √ | ? | √ | √ | √ | √ | √ | 8 |
| Tome J 2022 | √ | √ | √ | × | √ | × | √ | √ | √ | 7 |
| Witberg G 2021 | √ | √ | √ | √ | √ | √ | √ | √ | √ | 9 |
| Wong HL 2022 | √ | √ | √ | √ | √ | √ | √ | √ | √ | 9 |
| Hippisley-Co× J 2021 | √ | √ | √ | √ | √ | √ | √ | √ | √ | 9 |
| Jabagi MJ 2022 | √ | √ | √ | √ | √ | √ | √ | √ | √ | 9 |

√: Yes, ×: no, ?: unclear.

Table S3. Newcastle-Ottawa Scale for cohort studies.

| Study | Representativeness of the exposed cohort | Selection of the non-exposed cohort | Ascertainment of exposure | Demonstration that outcome of interest was not present at start of study | Comparability of cohorts on the basis of the design or analysis | Assessment of outcome | Was follow-up long enough for outcomes to occur | Adequacy of follow up of cohorts | Overall |
|---|---|---|---|---|---|---|---|---|---|
| Chou OHI 2022 | 1 | 1 | 1 | 1 | 2 | 1 | 1 | 1 | 9 |
| Chua GT 2021 | 1 | 1 | 1 | 1 | 2 | 1 | 1 | 1 | 9 |
| Dickerman BA 2022 | 1 | 1 | 1 | 1 | 2 | 1 | 1 | 1 | 9 |
| Farahmand R 2022 | 1 | 1 | 1 | 1 | 2 | 1 | 1 | 1 | 9 |
| lp S 2022 | 1 | 1 | 1 | 1 | 2 | 1 | 1 | 1 | 9 |
| Karlstad Ø 2022 | 1 | 1 | 1 | 1 | 2 | 1 | 1 | 1 | 9 |
| Lai FTT 2022a | 1 | 1 | 1 | 1 | 1 | 1 | 1 | 1 | 8 |
| Niesen MJM 2022 | 1 | 1 | 1 | 1 | 2 | 1 | 1 | 1 | 9 |
| Witberg G 2021 | 1 | 1 | 1 | 1 | 2 | 1 | 1 | 1 | 9 |
| Wong HL 2022 | 1 | 1 | 1 | 1 | 2 | 1 | 1 | 1 | 9 |
| Bardenheier BH 2021a | 1 | 1 | 1 | 1 | 2 | 1 | 1 | 1 | 9 |
| Bardenheier BH 2021b | 1 | 1 | 1 | 1 | 2 | 1 | 1 | 1 | 9 |
| Whiteley WN 2022 | 1 | 1 | 1 | 1 | 2 | 1 | 1 | 1 | 9 |

Table S4. Risk ratios of myocarditis after COVID-19 vaccination by dose number, vaccine type.

| | Studies, No. | Risk ratio (95%CI) | Heterogeneity, $I^2$ |
|---|---|---|---|
| First dose | | | |
| CoronaVac | 2 | 0.95(0.72-2.61) | 0.0% |
| ChAdOx1-S | 4 | 0.98(0.68-1.28) | 88.7% |
| mRNA-1273 | 2 | 1.33(0.81-1.86) | 0.0% |
| BNT162b2 | 7 | 1.12(0.83-1.42) | 81.9% |
| Second dose | | | |
| CoronaVac | 2 | 1.18(0.02-2.38) | 0.0% |
| ChAdOx1-S | 2 | 0.97(0.81-1.13) | 0.0% |
| mRNA-1273 | 4 | 7.27(5.33-9.21) | 21.7% |
| BNT162b2 | 6 | 1.55(1.36-1.74) | 23.0% |
